# Interventions to negate aggravated diarrhoeal disease due to climate change - A scoping review protocol

**DOI:** 10.1101/2025.06.03.25328790

**Authors:** Andres Madriz-Montero, Frederike Kooiman, Francis Ruiz, Fiammetta M. Bozzani

## Abstract

**Background:** Climate change is expected to magnify the risk of transmission of diarrhoeal disease, underscoring the need for effective and transformative adaptation. It is urgent to characterize the evidence of interventions that are available and delivered to mitigate the impact of climate change on diarrhoea to support decision making. The SPRINGS initiative has identified health technology assessment (HTA) as a potential approach to priority setting of planetary health interventions. This protocol describes the *a priori* objectives and methods to identify the adaptation interventions available to mitigate the impact of climate change on diarrhoeal disease and map the gaps between the available evidence and health technology assessment (HTA).

**Methods:** A search of nine bibliographic databases will be conducted by a library information professional with no time restriction to account for health, health economics and environmental science literature. Websites of major climate funds and stakeholders will be searched for grey and other literature. Two reviewers will independently screen titles and abstracts, and full texts based on predefined eligibility criteria. Data extraction will follow established climate change adaptation and HTA frameworks. Descriptive data analysis methods will be used for the synthesis of results through topic mapping, gap analysis and narrative synthesis.

**Ethics and dissemination:** Ethical approval to conduct the scoping review is not required as the information collected and reviewed is publicly available through database and website searches. Findings will be disseminated through peer-reviewed publications, workshops and conference presentations.

## Background and Rationale

Diarrhoea is currently the third leading cause of death in children under 5 years old and is responsible for almost 500 000 annual childhood deaths globally [1]. Current climate projections of increased precipitation and heat plus extreme weather events are expected to magnify the transmission of diarrhoeal disease [2]. It is estimated that climate change will cause an additional 48 000 annual deaths by 2030 in children aged under 15 years due to climate-sensitive diarrhoea [3], threatening decades of progress achieved in reducing the global burden [4].

The link between climate hazards and waterborne enteric disease transmission are well understood across high- and low-resource settings [5]. The impact of climate change on the burden of diarrhoeal disease depends on the dynamic interactions between climate hazards, vulnerabilities, pathogen exposures and risks [2]. In resource-constrained settings, added vulnerabilities stem from the insufficient progress in improved water supplies and sanitation [6], which limits their adaptive capacity to respond to the pressures of climate change [7].

Most of the literature on climate change and diarrhoea has predominantly focused on the climate drivers of disease, with limited attention to the interventions delivered to mitigate the health impact of diarrhoea [8]. A recent review of the effect of climate change adaptation on public health found that adaptation evaluations reporting on diarrhoea-related outcomes were mostly limited to safe-drinking water, sanitation, and hygiene (WASH) interventions [9]. Other research focused on strategies to prevent or manage diarrhoea have been extraneous to climate change [10], [11].

Despite the emphasis of adaptation on WASH, recent highly powered cluster trials found negligible benefits from among the most common types of WASH interventions used in low-and middle-income countries (LMICs) [12], [13], [14], underscoring the need for more effective interventions. Since individuals in resource-constrained settings are the most vulnerable to diarrhoeal health risk, it is necessary for countries to be able to prioritize the best climate-resilient strategies, including transformative adaptation for health in traditionally non-health sectors. To do so, it is urgent to characterize the evidence of interventions that are available and delivered to mitigate the impact of climate change on diarrhoea to support the priority setting of planetary health interventions.

Policy makers within the climate change space often lack structured appraisal processes and value assessments to support national level decision making [15]. The SPRINGS initiative [16], a global consortium that aims to improve policies and interventions to reduce the impact of climate change on diarrhoeal diseases, has identified health technology assessment (HTA) as a potential approach for priority setting of planetary health interventions. In two of SPRINGS’ case study countries (Ghana and Romania), the initiative will work with stakeholders and policy makers to develop a structured approach to assess, appraise and prioritize evidence-based interventions to mitigate climate-induced influences on diarrhoeal disease-related outcomes. As a first step, the process will define the full range of policy options available with a range of stakeholders across sectors.

To support this initial stage, this scoping review aims to identify the climate change adaptation strategies focused on diarrhoeal disease. In particular, we seek to map out the adaptation interventions, characterize them and summarize their available evidence in terms of their impact on health and non-health outcomes and other criteria relevant to HTA, to support topic selection and identify evidence gaps for the assessment stage of the HTA process proposed by SPRINGS.

## Methods

This scoping review protocol is informed by the preferred reporting items for systematic reviews and meta-analyses (PRISMA) guidelines [17]. Hence, we outline our methods *a priori* for this scoping review. This protocol will be registered online on the Open Science Framework repository.

We used a PICO framework to develop this scoping review protocol and to identify the research questions. The population (P) of interest includes climate-vulnerable individuals in all countries; the interventions (I) are adaptation interventions to reduce the impact of climate change on diarrhoeal disease; our aim will be to identify the evidence on diarrhoeal and other health and non-health outcomes (O), as well as other dimensions relevant to the HTA process; and we are interested in a broad range of empirical study designs (S).

### Stage 1: Identifying the research question and objectives

Two research questions were defined through consultation with the research team, within the broader SPRINGS initiative framework.

i. What adaptation interventions are delivered at any level (individual, community, regional, national) in response to climate change to negate aggravated diarrhoeal disease?
ii. What evidence is available about the interventions in terms of outcomes across sectors and other HTA-relevant dimensions that can support the priority setting process of planetary health interventions?

The overall objectives were defined for the scoping review framed by the research questions:

i. Identify and characterize the adaptation interventions that are delivered at any level in response to climate change to mitigate impact on diarrhoeal disease
ii. Map the available evidence and identify evidence gaps of HTA-relevant dimensions to support the priority setting of planetary health interventions

### Stage 2: Identifying the relevant literature

Systematic literature searches will be conducted in nine bibliographic databases to account for health, health economics and environmental science literature given the recent recommendation by Dinh et *al*. [18]: BIOSIS Citation Index, CINAHL Complete, Econlit, Embase Classic+Embase, Global Health, GreenFILE, Medline ALL, Scopus and Web of Science Core Collection. The search strategy was developed and tested in consultation with the team’s research librarian and will be conducted using both keywords and MeSH terms.

The search will focus on three subject headings, namely:

- Climate change AND
- Diarrhoeal disease AND
- Adaptation

The full search methodology is available in Appendix 1 and the full details of the search strings used for each database are available in Appendix 2, and published in the LSHTM Data Compass repository [19].

### Eligibility Criteria

| Criteria | Inclusion | Exclusion |
| --- | --- | --- |
| Publication Period | Any date | None |
| **Location** | Any country | None |
| **Language** | Language will be limited to English | Non-English language articles will be excluded in full text screening |
| **Study Type** | We will include the following types of study designs: randomized controlled studies, non-randomized and quasi-experimental designs, ecological studies, observational studies and systematic reviews and metanalysis study designs. Both quantitative and qualitative studies will be taken under consideration that report on outcomes | Articles that are letters to the editor, news articles, commentaries, and reviews other than systematic reviews. Articles where full text is not available |
| **Climate-sensitive diarrhoea pathogens** | Pathogens relevant to the SPRINGS initiative: Campylobacter, Giardia, Cryptosporidium and rotavirus<br>-All cause diarrhoea | Articles that report on exclusively on other pathogens |
| **Adaptation to Climate Change** | Adaptation strategies delivered to mitigate the impact of climate change on diarrhoea | Climate change mitigation<br>Articles where interventions are not described<br>Articles that do not evaluate the impact of an adaptation intervention or that don't report any outcomes<br>Articles that do not state the climate hazard that is addressed by the adaptation intervention |

Additional sources of literature from hand-searching relevant reference lists will be included. Grey an other published literature will be incorporated through review of the websites of the major climate funds including the Green Climate Fund (GCF), the Global Environment Facility (GEF), the Climate Investment Funds (CIF) and the Adaptation Fund, and stakeholders such as the Intergovernmental Panel on Climate Change (IPCC), the World Health Organization (WHO), the World Bank and Climate-ADAPT.

### Stage 3: Data management, screening and selection of eligible studies

After duplicate results are removed, the citations of bibliographic database and grey literature searches will be uploaded into Rayyan software [20] for reference management and screening. Eligibility criteria will be used to identify articles for inclusion as described below. All reviewers will be trained in the use of the screening software.

Before screening begins, the authors will test the article selection process by piloting the inclusion and exclusion criteria on a random sample of ten articles to ensure consistency and clarity of eligibility. If any issues arise, consensus on how to apply the criteria will be reached through discussion. After testing, all references will be initially screened independently for eligibility based on title and abstract relevance by two reviewers (AMM and FK). A sample of the excluded articles will be randomly selected for confirmation by another member of the research team (FR). Then, the full manuscripts allocated by Rayyan will be screened for inclusion and exclusion criteria by two independent reviewers (AMM and FK). Reasons for study exclusion will be documented at the full-text screening stage.

### Stage 4: Data extraction

After screening for eligibility, data from selected studies will be extracted into a data extraction table to provide a characterization of the interventions in line with the review’s objectives. As with the screening, the extraction table will be pre-tested against ten articles by the research team before extraction. All authors will extract the relevant information from the studies and thematically categorize them based on relevant adaptation frameworks [9], [21] and HTA reporting guidelines. Data will be extracted on the type of climate hazard evaluated (e.g. flood, extreme heat), the adaptation response type (e.g. infrastructure, technological); the intervention sector (e.g. water, forestry and agriculture); the level of implementation (national, regional, local); reported HTA-relevant outcomes (e.g. effectiveness, economic, ethical, legal), and decision making support dimensions. The data extraction table is provided in Appendix 3.

### Stage 5: Analysis and synthesis of results

This scoping review will follow the ROSES flow diagram [22] to showcase the review process used to achieve the final publication set. We will provide a comprehensive list of adaptation interventions thematically categorized and with their respective evidence. We will also use descriptive data analysis to showcase the results. We will produce heat maps and cross-tables of adaptation interventions across themes (HTA outcomes, adaptation response type, sector) for topic mapping and gap analysis, as well as narrative synthesis of all included studies by relevant themes that will support the identification of topics and the assessment stage of the HTA process for the adaptation strategies.

### Ethics and Dissemination

Ethical approval to conduct the scoping review is not required as the information collected and reviewed is publicly available through database and website searches.

This scoping review protocol will be shared with SPRINGS’ and country stakeholders and made available as an open-access report. Key findings of the scoping review will be disseminated through peer-reviewed publications, meetings and workshops, and conference presentations.

### Protocol Limitations

We foresee several limitations for the scoping review. First, as we have restricted the search to English language only, we acknowledge that there may be studies in other languages that specify relevant climate change adaptation interventions against diarrhoeal disease that are not captured by the search. In the same way, although we will not impose any geographical or time restrictions, we understand that the databases searched may not include all countries and relevant studies. To minimize this limitation, we will include gray and other published literature in our searches, however it will be constrained to websites of major funds and stakeholders and therefore may not be comprehensive. Due to resource limitations, we will only conduct a quality or bias assessment for the published literature to develop the evidence gap maps, and we acknowledge that findings may be influenced by limited reliability and quality and be subject to selection bias. This limitation is particularly important as further efforts will be required to assess risk if the evidence is used for a priority setting process.

## Data Availability

All data produced in the present work are contained in the manuscript

https://datacompass.lshtm.ac.uk/id/eprint/4464/

## Appendix 1

### Search methodology for “Interventions to negate aggravated diarrhoeal disease due to climate change - A scoping review”

### Methods

A draft search strategy was compiled in the Ovid Medline ALL database by an experienced information professional. The search strategy included strings of terms, synonyms and subject headings to reflect the following concepts: diarrhoea; climate change including floods, droughts, wildfires and heat; adaptation and risk. No limits were added to the search.

The search strategy was refined with the project team until the retrieved results reflected the scope of the project. Once agreed, the draft searches were adapted for each database to incorporate database-specific syntax and controlled vocabularies.

Search strategies were run on the databases listed in table 1 on 14 and 15 November 2024.

**Table 1.**
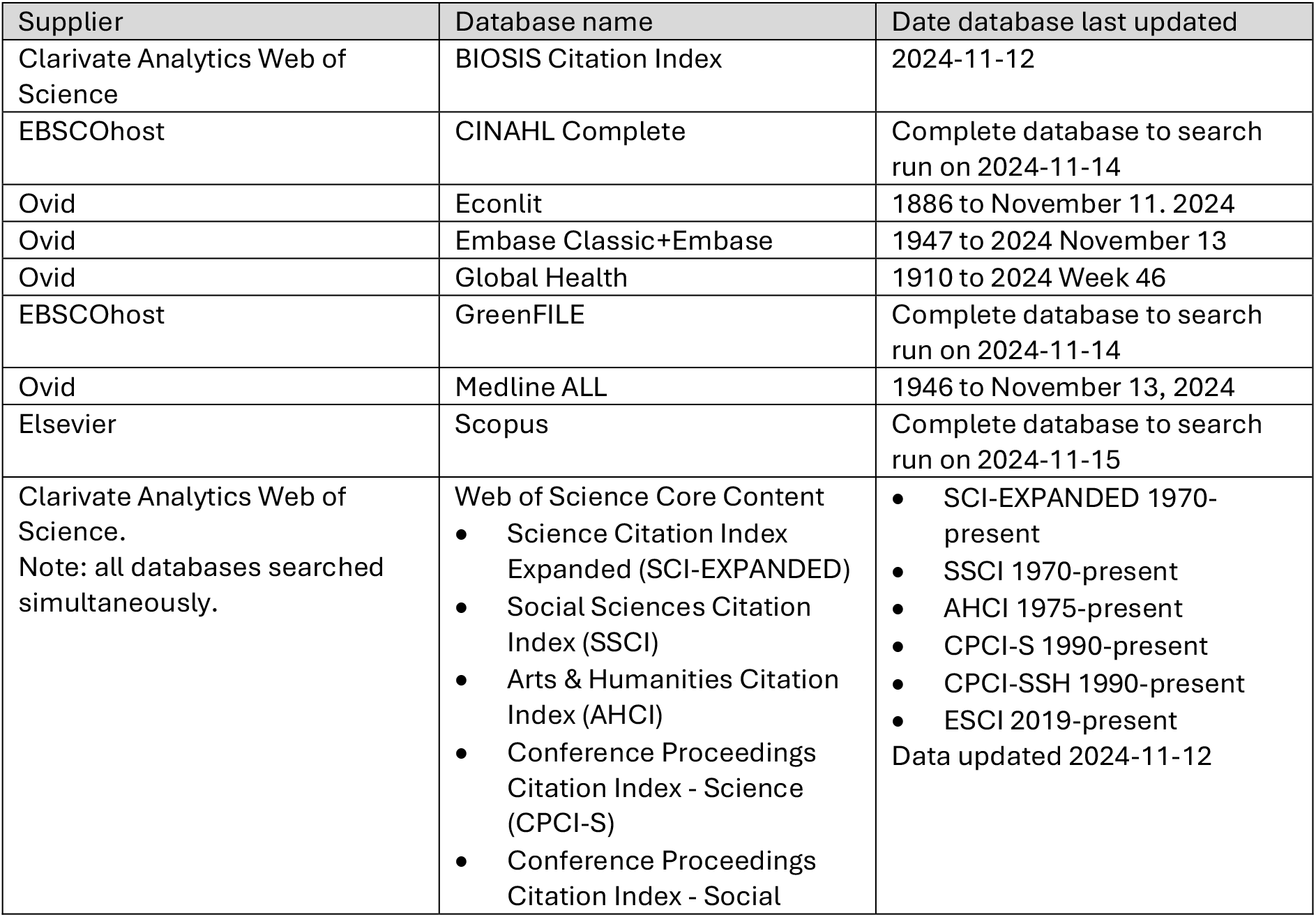
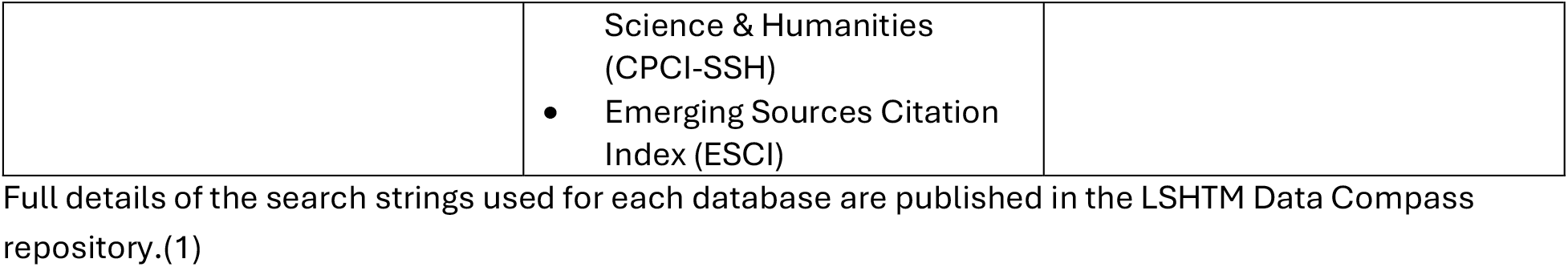
List of databases included in the search. Full details of the search strings used for each database are published in the LSHTM Data Compass repository.(1)

| Supplier | Database name | Date database last updated |
| --- | --- | --- |
| Clarivate Analytics Web of Science | BIOSIS Citation Index | 2024-11-12 |
| EBSCOhost | CINAHL Complete | Complete database to search run on 2024-11-14 |
| Ovid | Econlit | 1886 to November 11. 2024 |
| Ovid | Embase Classic+Embase | 1947 to 2024 November 13 |
| Ovid | Global Health | 1910 to 2024 Week 46 |
| EBSCOhost | GreenFILE | Complete database to search run on 2024-11-14 |
| Ovid | Medline ALL | 1946 to November 13, 2024 |
| Elsevier | Scopus | Complete database to search run on 2024-11-15 |
| Clarivate Analytics Web of Science.<br>Note: all databases searched simultaneously. | Web of Science Core Content<br>Science Citation Index Expanded (SCI-EXPANDED)<br>Social Sciences Citation Index (SSCI)<br>Arts & Humanities Citation Index (AHCI)<br>Conference Proceedings Citation Index - Science (CPCI-S)<br>Conference Proceedings Citation Index - Social | SCI-EXPANDED 1970-present<br>SSCI 1970-present<br>AHCI 1975-present<br>CPCI-S 1990-present<br>CPCI-SSH 1990-present<br>ESCI 2019-present<br>Data updated 2024-11-12 |
|  | Science & Humanities<br>(CPCI-SSH)<br>Emerging Sources Citation Index (ESCI) |  |

All results from the database searches were exported and uploaded to EndNote 21 software. Duplicates were identified and removed using the method described on the LAORS blog.(2) Deduplicated search results were exported from EndNote 21 in .ris format.

### Results

Details of results from each database are shown in table 2. A total of 6920 results were retrieved, 3996 duplicates were removed and 2924 were put forward for screening.

**Table 2.**
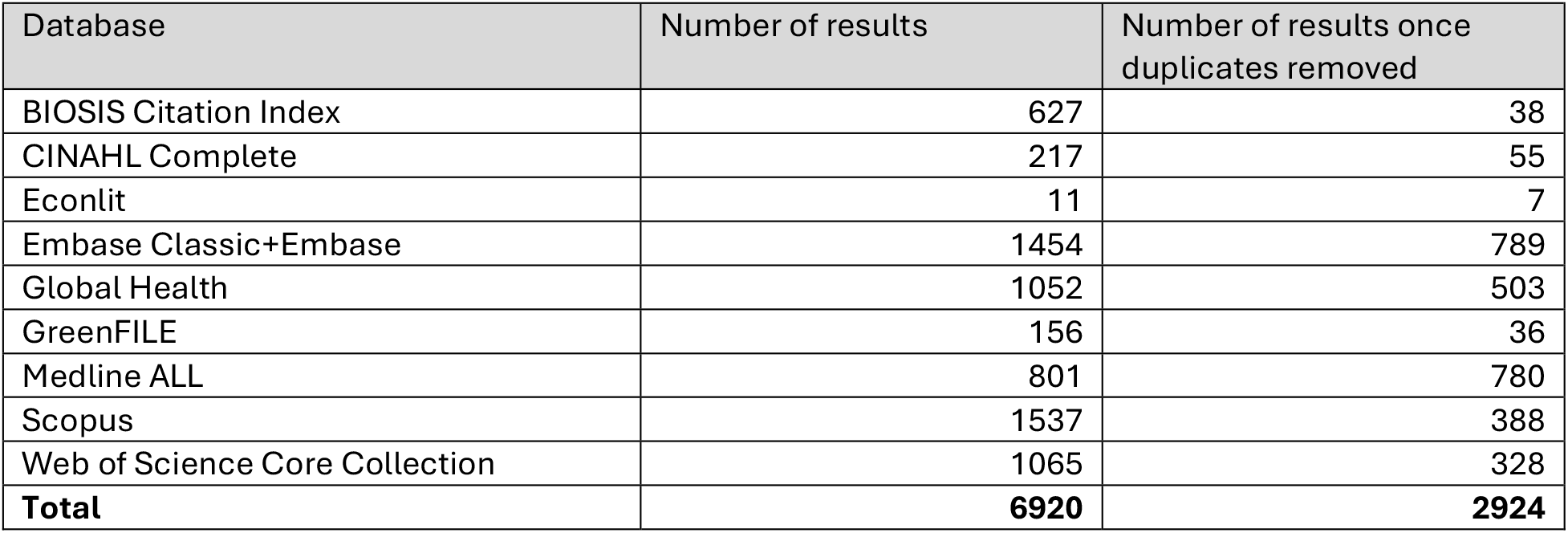
Search results retrieved from each database.

| Database | Number of results | Number of results once duplicates removed |
| --- | --- | --- |
| BIOSIS Citation Index | 627 | 38 |
| CINAHL Complete | 217 | 55 |
| Econlit | 11 | 7 |
| Embase Classic+Embase | 1454 | 789 |
| Global Health | 1052 | 503 |
| GreenFILE | 156 | 36 |
| Medline ALL | 801 | 780 |
| Scopus | 1537 | 388 |
| Web of Science Core Collection | 1065 | 328 |
| **Total** | **6920** | **2924** |

## Appendix 2: Full search strings

Interventions to negate aggravated diarrheal disease due to climate change - A scoping review: Final search strategies Jane Falconer, Francis Ruiz, Andres Madriz Montero

Search methods

A draft search strategy was compiled in the OvidSP Medline ALL database by an experienced information specialist. The search strategy included strings of terms and synonyms to reflect the following concepts:

Concept 1: diarrhoea

Concept 2: climate change, including floods, droughts, wildfires and heat

Concept 3: adaptation or risk.

No limits on language, publication date or study methodology was added to the search.

Search strategies were run on the databases listed below on 14 and 15 November 2024. Clarivate Analytics Web of Science BIOSIS Citation Index 2024-11-12

EBSCOhost CINAHL Complete Complete database to search run on 2024-11-14

Ovid Econlit 1886 to November 11. 2024

Ovid Embase Classic+Embase 1947 to 2024 November 13

Ovid Global Health 1910 to 2024 Week 46

EBSCOhost GreenFILEComplete database to search run on 2024-11-14 Ovid Medline ALL 1946 to November 13, 2024

Elsevier Scopus Complete database to search run on 2024-11-15

Clarivate Analytics Web of Science. Web of Science Core Content Data updated 2024-11-12

Science Citation Index Expanded (SCI-EXPANDED) SCI-EXPANDED 1970-present

Social Sciences Citation Index (SSCI) SSCI 1970-present

Arts & Humanities Citation Index (AHCI) AHCI 1975-present

Conference Proceedings Citation Index - Science (CPCI-S) CPCI-S 1990-present

Conference Proceedings Citation Index - Social Science & Humanities (CPCI-SSH) CPCI-SSH 1990-present

Emerging Sources Citation Index (ESCI) ESCI 2019-present

All results from the database searches were exported and uploaded to EndNote 21 software. Duplicates were identified and removed using the method described on the LAORS blog (Falconer, Jane, Removing duplicates from an EndNote library. Library & Archives Service Blog: London School of Hygiene & Tropical Medicine. 2018. [online blog] http://blogs.lshtm.ac.uk/library/2018/12/07/removing-duplicates-from-an-endnote-library/). Deduplicated search results were exported from EndNote 21 in .ris format.

Search strategies

This section provides full details of all search strings used for bibliographic databases, with dates and number of references returned and notes explaining any unusual search techniques or syntax.

Ovid Medline ALL

Database name Medline ALL Database platform Ovid

Dates of database coverage 1946 to November 13, 2024

Date searched 2024-11-14

Number of results 801

EndNote deduplication order 1

Results after duplications removed 780

Notes

Search lines ending in a ‘/’ are subject heading searches.

Search lines beginning ‘exp’ are exploded subject heading searches.

Two-letter codes at the end of search lines designate the fields to search. Fields codes used are: TI: title

AB: abstract

adjn searches for words within n words of each other.

or/x-y combines search sets in the range x-y with Boolean operator OR.

* is used for truncation of words.

? finds 0 or 1 character in that placeholder (optional wildcard).

words in square brackets […] are comments and are ignored.

1. climate change/ or global warming/ 35147
2. Greenhouse Gases/ or Greenhouse Effect/ 8839
3. Carbon footprint/ 1425
4. (exp Weather/ or exp Climate/) and exp Disasters/ 18365
5. (climat* adj3 (change? or changing or variab* or disrupt* or hazard* or impact* or vulnerab* or function* or extreme or catastroph* or emergenc* or shift* or dynamic* or sensitiv*)).ti,ab. 86252
6. extreme weather.ti,ab. 2708
7. (((global or planet* or world or climat* or earth or worldwide or world-wide) adj1 (heating or warming or temperature*)) or heat-dome).ti,ab. 21280
8. ((global or planet* or world or earth or worldwide or world-wide) adj2 (“co2” or carbon or methane or ozone or nitrous oxide)).ti,ab. 4925
9. ((greenhouse or green-house) adj2 (gas* or effect*)).ti,ab. 18063
10. ((“co2” or carbon or GHG or CH4 or methane or N2O or nitro* oxide* or non-methane volatile organic compound* or NMVOC* or sulphur dioxide* or ozone or O3 or fluorinated gase* or F-Gas* or hydrofluorocarbon* or HFCs or perfluorocarbon* or PFCs or sulfur hexafluoride* or sulphur hexafluoride* or SF6 or chlorofluorocarbon* or CFCs or hydrochlorofluorocarbon* or HCFCs or halos or air pollut* or aerosol* or ((short-lived or shortlived) adj3 pollutant*) or SLCP*) adj3 (mitigat* or reduc* or save* or saving or tackl* or address* or halt* or stop* or abat* or remission* or moderat* or modif* or control* or zero or diminish* or decreas* or eliminat* or elevat* or emission* or rise* or increas* or declin* or sink* or sequester* or neutral or offset or storage or store or storing or captur* or perform* or efficien* or conservation or remov*)).ti,ab. 136957
11. (emission* adj5 (elevat* or rise or increas* or decreas* or reduc* or offset*)).ti,ab. 36013
12. tipping point?.ti,ab. 1886
13. (carbon adj3 footprint?).ti,ab. 3298
14. (de-carbon or de-carboni* or decarbon or decarboni*).ti,ab. 1443
15. ((energy* or fuel*) adj3 (green or efficien* or reduc* or renewable* or save* or saving or clean* or transition*)).ti,ab. 75896
16. ((fossil fuel* or coal or oil or gas or carbon) adj3 (phas* out or exit* or transition* or transform* or clos* or shift* or decommission* or shut* down or displac* or low* or neutral* or sustainab*)).ti,ab. 32170
17. (net-zero or netzero).ti,ab. 1019
18. just transition*.ti,ab.57
19. BECCS.ti,ab. 60
20. petagram*.ti,ab. 103
21. wildfires/ or droughts/ or floods/ 19579
22. ((rain* or precipitation) adj4 (extreme or elevate? or elevating or irregular or intense or reduced or pattern* or anomal* or deficit* or surplus* or shortfall*)).ti,ab. 5980
23. (((fire? or burnt or inferno or conflagration) adj2 (change* or variab* or extreme or expand or expansion or intense or elevate? or increas*or catastroph* or emergenc* or shift* or dynamic* or frequen* or more or forest* or landscape? or wildland*)) or wildfire* or bushfire* or megafire*).ti,ab. 8098
24. (flood* or (water adj2 (stress or deficit or scarce or scarcit*)) or drought? or (climat* adj2 (wet* or dry*))).ti,ab. 70429
25. (habitat adj2 (loss or fragment* or disturb* or alter or altered or destruct*)).ti,ab. 6242
26. (deforestation or reforestation or “slash-and-burn” or desertificat* or (forest* adj2 (degradat* or conservat* or declin* or decreas*))).ti,ab. 7765
27. 27 or/1-26 420475
28. exp Diarrhea/ or (diarrhoea* or diarrhea*).ti,ab. 147422
29. ((faecal or fecal or faeces or feces or stool?) adj2 (consisten* or liquid* or water*)).ti,ab. 5756
30. Waterborne Diseases/ or ((water-borne or waterborne) adj1 (disease* or infect* or illness* or pathogen*)).ti,ab. 3023
31. Gastroenteritis/ or intestinal diseases, parasitic/ or (gastroenteritis or ((infect* or illness* or pathogen* or microbio* or bacteri* or virus* or viral or protozoa*) adj1 (intestin* or enteric* or gastro* or bowel))).ti,ab. 92059
32. Campylobacter Infections/ or exp Campylobacter/ or campylobacter*.ti,ab. 21511
33. giardiasis/ or exp Giardia/ or (giardiasis or giardiases or giardia).ti,ab. 12708
34. cryptosporidiosis/ or exp Cryptosporidium/ or (cryptosporidiosis or cryptosporidioses or cryptosporidium).ti,ab. 12126
35. Rotavirus Infections/ or Rotavirus/ or rotavirus.ti,ab. 17744 36 or/28-35 265015
36. exp Adaptation, Physiological/ or exp Adaptation, Biological/ or exp Adaptation, Psychological/ 305306
37. risk/ or risk management/ or risk assessment/ or risk adjustment/ or “risk evaluation and mitigation”/ 464062
38. (adaptation* or risk or vulnerable or vulnerabilit* or sustainab* or resilient or resilienc*).ti,ab. 3599621 40 or/37-39 4000091

41 27 and 36 and 40 803

42 remove duplicates from 41 801

Ovid Embase Classic+Embase

Database name Embase Classic+Embase Database platform Ovid

Dates of database coverage 1947 to 2024 November 13

Date searched 2024-11-14

Number of results 1454 EndNote deduplication order 2

Results after duplications removed 789

Notes

Search lines ending in a ‘/’ are subject heading searches.

Search lines beginning ‘exp’ are exploded subject heading searches.

Two-letter codes at the end of search lines designate the fields to search. Fields codes used are: TI: title

AB: abstract

adjn searches for words within n words of each other.

or/x-y combines search sets in the range x-y with Boolean operator OR.

* is used for truncation of words.

? finds 0 or 1 character in that placeholder (optional wildcard). words in square brackets […] are comments and are ignored.

1. exp climate change/ or exp environmental impact/ or exp environmental footprint/ or greenhouse gas/ or exp greenhouse gas emission/ 191639
2. (exp weather/ or exp climate/) and exp disaster/ 4459
3. (climat* adj3 (change? or changing or variab* or disrupt* or hazard* or impact* or vulnerab* or function* or extreme or catastroph* or emergenc* or shift* or dynamic* or sensitiv*)).ti,ab. 83016
4. extreme weather.ti,ab. 2676
5. (((global or planet* or world or climat* or earth or worldwide or world-wide) adj1 (heating or warming or temperature*)) or heat-dome).ti,ab. 21297
6. ((global or planet* or world or earth or worldwide or world-wide) adj2 (“co2” or carbon or methane or ozone or nitrous oxide)).ti,ab. 4907
7. ((greenhouse or green-house) adj2 (gas* or effect*)).ti,ab. 18759
8. ((“co2” or carbon or GHG or CH4 or methane or N2O or nitro* oxide* or non-methane volatile organic compound* or NMVOC* or sulphur dioxide* or ozone or O3 or fluorinated gase* or F-Gas* or hydrofluorocarbon* or HFCs or perfluorocarbon* or PFCs or sulfur hexafluoride* or sulphur hexafluoride* or SF6 or chlorofluorocarbon* or CFCs or hydrochlorofluorocarbon* or HCFCs or halos or air pollut* or aerosol* or ((short-lived or shortlived) adj3 pollutant*) or SLCP*) adj3 (mitigat* or reduc* or save* or saving or tackl* or address* or halt* or stop* or abat* or remission* or moderat* or modif* or control* or zero or diminish* or decreas* or eliminat* or elevat* or emission* or rise* or increas* or declin* or sink* or sequester* or neutral or offset or storage or store or storing or captur* or perform* or efficien* or conservation or remov*)).ti,ab. 154000
9. (emission* adj5 (elevat* or rise or increas* or decreas* or reduc* or offset*)).ti,ab. 39705
10. tipping point?.ti,ab. 2092
11. (carbon adj3 footprint?).ti,ab. 3849
12. (de-carbon or de-carboni* or decarbon or decarboni*).ti,ab. 1232
13. ((energy* or fuel*) adj3 (green or efficien* or reduc* or renewable* or save* or saving or clean* or transition*)).ti,ab. 74420
14. ((fossil fuel* or coal or oil or gas or carbon) adj3 (phas* out or exit* or transition* or transform* or clos* or shift* or decommission* or shut* down or displac* or low* or neutral* or sustainab*)).ti,ab. 33031
15. (net-zero or netzero).ti,ab. 1001
16. just transition*.ti,ab.53
17. BECCS.ti,ab. 62
18. petagram*.ti,ab. 99
19. environmental impact/ or drought/ or flooding/ or waterlogging/ or exp wildfire/ 70622
20. ((rain* or precipitation) adj4 (extreme or elevate? or elevating or irregular or intense or reduced or pattern* or anomal* or deficit* or surplus* or shortfall*)).ti,ab. 5900
21. (((fire? or burnt or inferno or conflagration) adj2 (change* or variab* or extreme or expand or expansion or intense or elevate? or increas*or catastroph* or emergenc* or shift* or dynamic* or frequen* or more or forest* or landscape? or wildland*)) or wildfire* or bushfire* or megafire*).ti,ab. 8751
22. (flood* or (water adj2 (stress or deficit or scarce or scarcit*)) or drought? or (climat* adj2 (wet* or dry*))).ti,ab. 68221
23. deforestation/ 3220
24. (habitat adj2 (loss or fragment* or disturb* or alter or altered or destruct*)).ti,ab. 5675
25. (deforestation or reforestation or “slash-and-burn” or desertificat* or (forest* adj2 (degradat* or conservat* or declin* or decreas*))).ti,ab. 7740

26 or/1-25 513412

1. exp Diarrhea/ or (diarrhoea* or diarrhea*).ti,ab. 401302
2. ((faecal or fecal or faeces or feces or stool?) adj2 (consisten* or liquid* or water*)).ti,ab. 9300
3. water borne disease/ or ((water-borne or waterborne) adj1 (disease* or infect* or illness* or pathogen*)).ti,ab. 4006
4. gastroenteritis/ 28760
5. intestine infection/ 14256
6. gastroenteritis/ or intestine infection/ or (gastroenteritis or ((infect* or illness* or pathogen* or microbio* or bacteri* or virus* or viral or protozoa*) adj1 (intestin* or enteric* or gastro* or bowel))).ti,ab. 121252
7. exp Campylobacter/ or exp campylobacteriosis/ or campylobacter*.ti,ab.29126
8. giardiasis/ or exp Giardia/ or (giardiasis or giardiases or giardia).ti,ab. 17633
9. cryptosporidiosis/ or exp Cryptosporidium/ or (cryptosporidiosis or cryptosporidioses or cryptosporidium).ti,ab. 16751
10. exp Rotavirus/ or Rotavirus infection/ or rotavirus.ti,ab. 23788

37 or/27-36 551277

1. exp adaptation/ 237283
2. risk/ or attributable risk/ or compound risk/ or exp environmental risk/ or infection risk/ or population risk/ or exp risk assessment/ or exp risk attitude/ or exp risk behavior/ or risk benefit analysis/ or “risk evaluation and mitigation strategy”/ or exp risk management/ or risk perception/ or risk reduction/ or exp vulnerability/ 1662959
3. environmental resilience/ or climate resilience/ 1324
4. (adaptation* or risk or vulnerable or vulnerabilit* or sustainab* or resilient or resilienc*).ti,ab. 5076043 42 or/38-41 5628142

43 26 and 37 and 42 1471

44 remove duplicates from 43 1454

Ovid Global Health

Database name Global Health Database platform Ovid

Dates of database coverage 1910 to 2024 Week 46

Date searched 2024-11-14

Number of results 1052

EndNote deduplication order 3

Results after duplications removed 503

Notes

Search lines ending in a ‘/’ are subject heading searches.

Search lines beginning ‘exp’ are exploded subject heading searches.

Two-letter codes at the end of search lines designate the fields to search. Fields codes used are: TI: title

AB: abstract

adjn searches for words within n words of each other.

or/x-y combines search sets in the range x-y with Boolean operator OR.

* is used for truncation of words.

? finds 0 or 1 character in that placeholder (optional wildcard). words in square brackets […] are comments and are ignored.

1. exp climate change/ 16197
2. greenhouse gases/ or carbon dioxide/ or greenhouse effect/ or methane/ or nitrogen oxides/ 19818
3. exp environmental impact/ 15433
4. (exp weather/ or exp climate/) and exp disasters/ 654
5. (climat* adj3 (change? or changing or variab* or disrupt* or hazard* or impact* or vulnerab* or function* or extreme or catastroph* or emergenc* or shift* or dynamic* or sensitiv*)).ti,ab. 21929
6. extreme weather.ti,ab. 1073
7. (((global or planet* or world or climat* or earth or worldwide or world-wide) adj1 (heating or warming or temperature*)) or heat-dome).ti,ab. 4018
8. ((global or planet* or world or earth or worldwide or world-wide) adj2 (“co2” or carbon or methane or ozone or nitrous oxide)).ti,ab. 313
9. ((greenhouse or green-house) adj2 (gas* or effect*)).ti,ab. 4540
10. ((“co2” or carbon or GHG or CH4 or methane or N2O or nitro* oxide* or non-methane volatile organic compound* or NMVOC* or sulphur dioxide* or ozone or O3 or fluorinated gase* or F-Gas* or hydrofluorocarbon* or HFCs or perfluorocarbon* or PFCs or sulfur hexafluoride* or sulphur hexafluoride* or SF6 or chlorofluorocarbon* or CFCs or hydrochlorofluorocarbon* or HCFCs or halos or air pollut* or aerosol* or ((short-lived or shortlived) adj3 pollutant*) or SLCP*) adj3 (mitigat* or reduc* or save* or saving or tackl* or address* or halt* or stop* or abat* or remission* or moderat* or modif* or control* or zero or diminish* or decreas* or eliminat* or elevat* or emission* or rise* or increas* or declin* or sink* or sequester* or neutral or offset or storage or store or storing or captur* or perform* or efficien* or conservation or remov*)).ti,ab. 22776
11. (emission* adj5 (elevat* or rise or increas* or decreas* or reduc* or offset*)).ti,ab. 6491
12. tipping point?.ti,ab. 182
13. (carbon adj3 footprint?).ti,ab. 1040
14. (de-carbon or de-carboni* or decarbon or decarboni*).ti,ab. 175
15. ((energy* or fuel*) adj3 (green or efficien* or reduc* or renewable* or save* or saving or clean* or transition*)).ti,ab. 12869
16. ((fossil fuel* or coal or oil or gas or carbon) adj3 (phas* out or exit* or transition* or transform* or clos* or shift* or decommission* or shut* down or displac* or low* or neutral* or sustainab*)).ti,ab. 6806
17. sustainability/ or “sustainable land use”/ 19507
18. (net-zero or netzero).ti,ab. 115
19. just transition*.ti,ab.17
20. BECCS.ti,ab. 8
21. petagram*.ti,ab. 3
22. wildfires/ or drought/ or dry conditions/ or floods/ or flooded land/ 6566
23. deforestation/ or desertification/ 1244
24. ((rain* or precipitation) adj4 (extreme or elevate? or elevating or irregular or intense or reduced or pattern* or anomal* or deficit* or surplus* or shortfall*)).ti,ab. 1663
25. (((fire? or burnt or inferno or conflagration) adj2 (change* or variab* or extreme or expand or expansion or intense or elevate? or increas*or catastroph* or emergenc* or shift* or dynamic* or frequen* or more or forest* or landscape? or wildland*)) or wildfire* or bushfire* or megafire*).ti,ab. 1945
26. (flood* or (water adj2 (stress or deficit or scarce or scarcit*)) or drought? or (climat* adj2 (wet* or dry*))).ti,ab. 18952
27. (habitat adj2 (loss or fragment* or disturb* or alter or altered or destruct*)).ti,ab. 808
28. (deforestation or reforestation or “slash-and-burn” or desertificat* or (forest* adj2 (degradat* or conservat* or declin* or decreas*))).ti,ab. 2519

29 or/1-28 123205

1. exp diarrhoea/ or (diarrhoea* or diarrhea*).ti,ab. 73527
2. ((faecal or fecal or faeces or feces or stool?) adj2 (consisten* or liquid* or water*)).ti,ab. 3392
3. waterborne diseases/ or ((water-borne or waterborne) adj1 (disease* or infect* or illness* or pathogen*)).ti,ab. 7693
4. gastroenteritis/ or (gastroenteritis or ((infect* or illness* or pathogen* or microbio* or bacteri* or virus* or viral or protozoa*) adj1 (intestin* or enteric* or gastro* or bowel))).ti,ab. 47030
5. exp campylobacter/ or campylobacter*.ti,ab. 15090
6. exp giardia/ or (giardiasis or giardiases or giardia).ti,ab. 15993
7. exp cryptosporidium/ 12558
8. exp cryptosporidium/ or (cryptosporidiosis or cryptosporidioses or cryptosporidium).ti,ab. 13045
9. exp rotavirus/ or rotavirus.ti,ab.10823 39 or/30-38 146071
10. adaptation/ or acclimatization/ or adaptability/ 5274
11. risk/ or risk analysis/ or risk management/ or risk assessment/ or risk reduction/ 229281
12. (adaptation* or risk or vulnerable or vulnerabilit* or sustainab* or resilient or resilienc*).ti,ab. 887601 43 or/40-42 922996

44 29 and 39 and 43 1059

45 remove duplicates from 44 1052

Ovid Econlit

Database name Econlit Database platform Ovid

Dates of database coverage 1886 to November 11, 2024

Date searched 2024-11-14

Number of results 11

EndNote deduplication order 4

Results after duplications removed 7

Notes

Two-letter codes at the end of search lines designate the fields to search. Fields codes used are: TI: title

AB: abstract

adjn searches for words within n words of each other.

or/x-y combines search sets in the range x-y with Boolean operator OR.

* is used for truncation of words.

? finds 0 or 1 character in that placeholder (optional wildcard). words in square brackets […] are comments and are ignored.

1. (climat* adj3 (change? or changing or variab* or disrupt* or hazard* or impact* or vulnerab* or function* or extreme or catastroph* or emergenc* or shift* or dynamic* or sensitiv*)).ti,ab. 17956
2. extreme weather.ti,ab. 554
3. (((global or planet* or world or climat* or earth or worldwide or world-wide) adj1 (heating or warming or temperature*)) or heat-dome).ti,ab. 2821
4. ((global or planet* or world or earth or worldwide or world-wide) adj2 (“co2” or carbon or methane or ozone or nitrous oxide)).ti,ab. 801
5. ((greenhouse or green-house) adj2 (gas* or effect*)).ti,ab. 6256
6. ((“co2” or carbon or GHG or CH4 or methane or N2O or nitro* oxide* or non-methane volatile organic compound* or NMVOC* or sulphur dioxide* or ozone or O3 or fluorinated gase* or F-Gas* or hydrofluorocarbon* or HFCs or perfluorocarbon* or PFCs or sulfur hexafluoride* or sulphur hexafluoride* or SF6 or chlorofluorocarbon* or CFCs or hydrochlorofluorocarbon* or HCFCs or halos or air pollut* or aerosol* or ((short-lived or shortlived) adj3 pollutant*) or SLCP*) adj3 (mitigat* or reduc* or save* or saving or tackl* or address* or halt* or stop* or abat* or remission* or moderat* or modif* or control* or zero or diminish* or decreas* or eliminat* or elevat* or emission* or rise* or increas* or declin* or sink* or sequester* or neutral or offset or storage or store or storing or captur* or perform* or efficien* or conservation or remov*)).ti,ab. 15877
7. (emission* adj5 (elevat* or rise or increas* or decreas* or reduc* or offset*)).ti,ab. 11557
8. tipping point?.ti,ab. 416
9. (carbon adj3 footprint?).ti,ab. 569
10. (de-carbon or de-carboni* or decarbon or decarboni*).ti,ab. 1780
11. ((energy* or fuel*) adj3 (green or efficien* or reduc* or renewable* or save* or saving or clean* or transition*)).ti,ab. 17672
12. ((fossil fuel* or coal or oil or gas or carbon) adj3 (phas* out or exit* or transition* or transform* or clos* or shift* or decommission* or shut* down or displac* or low* or neutral* or sustainab*)).ti,ab. 6134
13. (net-zero or netzero).ti,ab. 502
14. just transition*.ti,ab.214
15. BECCS.ti,ab. 35
16. petagram*.ti,ab. 2
17. ((rain* or precipitation) adj4 (extreme or elevate? or elevating or irregular or intense or reduced or pattern* or anomal* or deficit* or surplus* or shortfall*)).ti,ab. 293
18. (((fire? or burnt or inferno or conflagration) adj2 (change* or variab* or extreme or expand or expansion or intense or elevate? or increas*or catastroph* or emergenc* or shift* or dynamic* or frequen* or more or forest* or landscape? or wildland*)) or wildfire* or bushfire* or megafire*).ti,ab. 574
19. (flood* or (water adj2 (stress or deficit or scarce or scarcit*)) or drought? or (climat* adj2 (wet* or dry*))).ti,ab. 4862
20. (habitat adj2 (loss or fragment* or disturb* or alter or altered or destruct*)).ti,ab. 99
21. (deforestation or reforestation or “slash-and-burn” or desertificat* or (forest* adj2 (degradat* or conservat* or declin* or decreas*))).ti,ab. 2404

22 or/1-21 57396

1. (diarrhoea* or diarrhea*).ti,ab. 283
2. ((faecal or fecal or faeces or feces or stool?) adj2 (consisten* or liquid* or water*)).ti,ab. 1
3. ((water-borne or waterborne) adj1 (disease* or infect* or illness* or pathogen*)).ti,ab. 73
4. (gastroenteritis or ((infect* or illness* or pathogen* or microbio* or bacteri* or virus* or viral or protozoa*) adj1 (intestin* or enteric* or gastro* or bowel))).ti,ab. 22
5. campylobacter*.ti,ab. 13
6. (giardiasis or giardiases or giardia).ti,ab. 2
7. (cryptosporidiosis or cryptosporidioses or cryptosporidium).ti,ab. 1
8. rotavirus.ti,ab. 17

31 or/23-30 399

32 (adaptation* or risk or vulnerable or vulnerabilit* or sustainab* or resilient or resilienc*).ti,ab. 196108 33 22 and 31 and 32 11

34 remove duplicates from 33 11

EBSCOhost CINAHL Complete

Database name CINAHL Complete Database platform EBSCOhost

Dates of database coverage Complete database to search date Date searched 2024-11-14

Number of results 217

EndNote deduplication order 5

Results after duplications removed 55

Notes

Two-letter codes at the start of search lines designate the fields to search. Fields codes used are: MH: subject heading

TI: title

AB: abstract

Nn searches for words within n words of each other.

* is used for truncation of words.

\# finds 0 or 1 character in that placeholder (optional wildcard).

S1 (MH “Climate Change+”) OR (MH “Greenhouse Effect”) 5,183 S2 (MH “Greenhouse Gases”) 378

S3 (MH “Carbon Footprint”) OR (MH “Conservation of Natural Resources+”) OR (MH “Environmental Sustainability”) 6,325 S4 (AB(climat* N4 (change# or changing or variab* or disrupt* or hazard* or impact* or vulnerab* or function* or extreme or

catastroph* or emergenc* or shift* or dynamic* or sensitiv*))) OR (TI(climat* N4 (change# or changing or variab* or disrupt* or hazard* or impact* or vulnerab* or function* or extreme or catastroph* or emergenc* or shift* or dynamic* or sensitiv*))) 7,691

S5 (AB “extreme weather”) OR (TI “extreme weather”) 415

S6 (AB(((global or planet* or world or climat* or earth or worldwide or world-wide) N2 (heating or warming or temperature*)) or heat-dome)) OR (TI(((global or planet* or world or climat* or earth or worldwide or world-wide) N2 (heating or warming or temperature*)) or heat-dome)) 1,203

S7 (AB((global or planet* or world or earth or worldwide or world-wide) N3 (“co2” or carbon or methane or ozone or “nitrous oxide”))) OR (TI((global or planet* or world or earth or worldwide or world-wide) N3 (“co2” or carbon or methane or ozone or “nitrous oxide”))) 139

S8 (AB((greenhouse or green-house) N3 (gas* or effect*))) OR (TI((greenhouse or green-house) N3 (gas* or effect*))) 783

S9 (AB((“co2” or carbon or GHG or CH4 or methane or N2O or “nitro* oxide*” or “non-methane volatile organic compound*” or NMVOC* or “sulphur dioxide*” or ozone or O3 or “fluorinated gase*” or F-Gas* or hydrofluorocarbon* or HFCs or perfluorocarbon* or PFCs or “sulfur hexafluoride*” or “sulphur hexafluoride*” or SF6 or chlorofluorocarbon* or CFCs or hydrochlorofluorocarbon* or HCFCs or halos or “air pollut*” or aerosol* or ((“short-lived” or shortlived) N4 pollutant*) or SLCP*) N4 (mitigat* or reduc* or save* or saving or tackl* or address* or halt* or stop* or abat* or remission* or moderat* or modif* or control* or zero or diminish* or decreas* or eliminat* or elevat* or emission* or rise* or increas* or declin* or sink* or sequester* or neutral or offset or storage or store or storing or captur* or perform* or efficien* or conservation or remov*))) OR (TI((“co2” or carbon or GHG or CH4 or methane or N2O or “nitro* oxide*” or “non-methane volatile organic compound*” or NMVOC* or “sulphur dioxide*” or ozone or O3 or “fluorinated gase*” or F-Gas* or hydrofluorocarbon* or HFCs or perfluorocarbon* or PFCs or “sulfur hexafluoride*” or “sulphur hexafluoride*” or SF6 or chlorofluorocarbon* or CFCs or hydrochlorofluorocarbon* or HCFCs or halos or “air pollut*” or aerosol* or ((“short-lived” or shortlived) N4 pollutant*) or SLCP*) N4 (mitigat* or reduc* or save* or saving or tackl* or address* or halt* or stop* or abat* or remission* or moderat* or modif* or control* or zero or diminish* or decreas* or eliminat* or elevat* or emission* or rise* or increas* or declin* or sink* or sequester* or neutral or offset or storage or store or storing or captur* or perform* or efficien* or conservation or remov*)))

8,045

S10 (AB(emission* N6 (elevat* or rise or increas* or decreas* or reduc* or offset*))) OR (TI(emission* N6 (elevat* or rise or increas* or decreas* or reduc* or offset*))) 1,538

S11 (TI “tipping point#”) or (AB “tipping point#”) 577

S12 (TI (carbon N4 footprint#)) or (AB (carbon N4 footprint#)) 446

S13 (AB(de-carbon or de-carboni* or decarbon or decarboni*)) OR (TI(de-carbon or de-carboni* or decarbon or decarboni*)) 82

S14 (AB((energy* or fuel*) N4 (green or efficien* or reduc* or renewable* or save* or saving or clean* or transition*))) OR (TI((energy* or fuel*) N4 (green or efficien* or reduc* or renewable* or save* or saving or clean* or transition*))) 4,796

S15 (AB((“fossil fuel*” or coal or oil or gas or carbon) N4 (“phas* out” or exit* or transition* or transform* or clos* or shift* or decommission* or “shut* down” or displac* or low* or neutral* or sustainab*))) OR (TI((“fossil fuel*” or coal or oil or gas or carbon) N4 (“phas* out” or exit* or transition* or transform* or clos* or shift* or decommission* or “shut* down” or displac* or low* or neutral* or sustainab*))) 2,380

S16 (TI (net-zero or netzero)) OR (AB (net-zero or netzero)) 126

S17 (TI “just transition*”) OR (AB “just transition*”) 19

S18 (TI BECCS) OR (AB BECCS) 9

S19 (TI petagram*) or (AB petagram*) 1

S20 (MH “Wildfires”) 336

S21 (MH “Natural Disasters”) 12,799

S22 (TI ((rain* or precipitation) N5 (extreme or elevate# or elevating or irregular or intense or reduced or pattern* or anomal* or deficit* or surplus* or shortfall*))) or (AB ((rain* or precipitation) N5 (extreme or elevate# or elevating or irregular or intense or reduced or pattern* or anomal* or deficit* or surplus* or shortfall*))) 199

S23 (TI (((fire# or burnt or inferno or conflagration) N3 (change* or variab* or extreme or expand or expansion or intense or elevate# or increas*or catastroph* or emergenc* or shift* or dynamic* or frequen* or more or forest* or landscape# or wildland*)) or wildfire* or bushfire* or megafire*) or (AB (((fire# or burnt or inferno or conflagration) N3 (change* or variab* or extreme or expand or expansion or intense or elevate# or increas*or catastroph* or emergenc* or shift* or dynamic* or frequen* or more or forest* or landscape# or wildland*)) or wildfire* or bushfire* or megafire*) 1,869

S24 (TI (flood* or (water N3 (stress or deficit or scarce or scarcit*)) or drought# or (climat* N3 (wet* or dry*)))) or (AB (flood* or (water N3 (stress or deficit or scarce or scarcit*)) or drought# or (climat* N3 (wet* or dry*)))) 3,634

S25 (TI (habitat N3 (loss or fragment* or disturb* or alter or altered or destruct*))) or (AB (habitat N3 (loss or fragment* or disturb* or alter or altered or destruct*))) 109

S26 (TI (deforestation or reforestation or “slash-and-burn” or desertificat* or (forest* adj2 (degradat* or conservat* or declin* or decreas*)))) or (AB (deforestation or reforestation or “slash-and-burn” or desertificat* or (forest* adj2 (degradat* or conservat* or declin* or decreas*)))) 277

S27 S1 OR S2 OR S3 OR S4 OR S5 OR S6 OR S7 OR S8 OR S9 OR S10 OR S11 OR S12 OR S13 OR S14 OR S15 OR S16 OR S17 OR S18 OR S19 OR S20 OR S21 OR S22 OR S23 OR S24 OR S25 OR S26 47,914

S28 (MH “Diarrhea”) 12,625

S29 (TI (diarrhoea* or diarrhea*)) or (AB (diarrhoea* or diarrhea*)) 21,449

S30 (TI ((water-borne or waterborne) N2 (disease* or infect* or illness* or pathogen*))) or (AB ((water-borne or waterborne) N2 (disease* or infect* or illness* or pathogen*))) 486

S31 (MH “Gastroenteritis”) 3,171

S32 (MH “Intestinal Diseases, Parasitic”) 775

S33 (TI (gastroenteritis or ((infect* or illness* or pathogen* or microbio* or bacteri* or virus* or viral or protozoa*) N2 (intestin* or enteric* or gastro* or bowel)))) or (AB (gastroenteritis or ((infect* or illness* or pathogen* or microbio* or bacteri* or virus* or viral or protozoa*) N2 (intestin* or enteric* or gastro* or bowel)))) 12,267

S34 (MH “Campylobacter”) 786

S35 (MH “Campylobacter Infections”) 879

S36 (TI campylobacter*) or (AB campylobacter*) 1,431 S37 (MH “Giardiasis”) 479

S38 (MH “Giardia Lamblia”) 204

S39 (TI (giardiasis or giardiases or giardia)) or (AB (giardiasis or giardiases or giardia)) 827

S40 (MH “Cryptosporidiosis”) 567

S41 (MH “Cryptosporidium”) 437

S42 (TI (cryptosporidiosis or cryptosporidioses or cryptosporidium)) or (AB (cryptosporidiosis or cryptosporidioses or cryptosporidium)) 851

S43 (MH “Rotaviruses”) OR (MH “Rotavirus Infections”) 1,766 S44 (TI rotavirus) or (AB rotavirus) 2,250

S45 S28 OR S29 OR S30 OR S31 OR S32 OR S33 OR S34 OR S35 OR S36 OR S37 OR S38 OR S39 OR S40 OR S41 OR S42 OR S43 OR S44 40,647

S46 (MH “Risk Assessment”) 176,097 S47 (MH “Risk Management”) 16,707 S48 (MH “Attributable Risk”) 569

S49 (MH “Adaptation, Physiological+”) 9,648 S50 (MH “Adaptation, Psychological”) 37,008 S51 (MH “Vulnerability”) 7,446

S52 (AB (adaptation* or risk or vulnerable or vulnerabilit* or sustainab* or resilient or resilienc*)) or (TI (adaptation* or risk or vulnerable or vulnerabilit* or sustainab* or resilient or resilienc*)) 1,059,573

S53 S46 OR S47 OR S48 OR S49 OR S50 OR S51 OR S52 1,168,386 S54 S27 AND S45 AND S53 217

EBSCOhost GreenFILE

Database name GreenFILE Database platform EBSCOhost

Dates of database coverage Complete database to search date Date searched 2024-11-14

Number of results 156

EndNote deduplication order 6

Results after duplications removed 36

Notes

Two-letter codes at the start of search lines designate the fields to search. Fields codes used are: TI: title

AB: abstract

Nn searches for words within n words of each other.

* is used for truncation of words.

\# finds 0 or 1 character in that placeholder (optional wildcard).

S1 (MH “Climate Change+”) OR (MH “Greenhouse Effect”) 1,956 S2 (MH “Greenhouse Gases”) 12,551

S3 (MH “Carbon Footprint”) OR (MH “Conservation of Natural Resources+”) OR (MH “Environmental Sustainability”) 1,414 S4 (AB(climat* N4 (change# or changing or variab* or disrupt* or hazard* or impact* or vulnerab* or function* or extreme or

catastroph* or emergenc* or shift* or dynamic* or sensitiv*))) OR (TI(climat* N4 (change# or changing or variab* or disrupt* or hazard* or impact* or vulnerab* or function* or extreme or catastroph* or emergenc* or shift* or dynamic* or sensitiv*))) 75,212

S5 (AB “extreme weather”) OR (TI “extreme weather”) 1,486

S6 (AB(((global or planet* or world or climat* or earth or worldwide or world-wide) N2 (heating or warming or temperature*)) or heat-dome)) OR (TI(((global or planet* or world or climat* or earth or worldwide or world-wide) N2 (heating or warming or temperature*)) or heat-dome)) 21,719

S7 (AB((global or planet* or world or earth or worldwide or world-wide) N3 (“co2” or carbon or methane or ozone or “nitrous oxide”))) OR (TI((global or planet* or world or earth or worldwide or world-wide) N3 (“co2” or carbon or methane or ozone or “nitrous oxide”))) 5,663

S8 (AB((greenhouse or green-house) N3 (gas* or effect*))) OR (TI((greenhouse or green-house) N3 (gas* or effect*))) 25,952 S9 (AB((“co2” or carbon or GHG or CH4 or methane or N2O or “nitro* oxide*” or “non-methane volatile organic compound*” or NMVOC* or “sulphur dioxide*” or ozone or O3 or “fluorinated gase*” or F-Gas* or hydrofluorocarbon* or HFCs or perfluorocarbon* or PFCs or “sulfur hexafluoride*” or “sulphur hexafluoride*” or SF6 or chlorofluorocarbon* or CFCs or hydrochlorofluorocarbon* or HCFCs or halos or “air pollut*” or aerosol* or ((“short-lived” or shortlived) N4 pollutant*) or SLCP*) N4 (mitigat* or reduc* or save* or saving or tackl* or address* or halt* or stop* or abat* or remission* or moderat* or modif* or control* or zero or diminish* or decreas* or eliminat* or elevat* or emission* or rise* or increas* or declin* or sink* or sequester* or neutral or offset or storage or store or storing or captur* or perform* or efficien* or conservation or remov*))) OR (TI((“co2” or carbon or GHG or CH4 or methane or N2O or “nitro* oxide*” or “non-methane volatile organic compound*” or NMVOC* or “sulphur dioxide*” or ozone or O3 or “fluorinated gase*” or F-Gas* or hydrofluorocarbon* or HFCs or perfluorocarbon* or PFCs or “sulfur hexafluoride*” or “sulphur hexafluoride*” or SF6 or chlorofluorocarbon* or CFCs or hydrochlorofluorocarbon* or HCFCs or halos or “air pollut*” or aerosol* or ((“short-lived” or shortlived) N4 pollutant*) or SLCP*) N4 (mitigat* or reduc* or save* or saving or tackl* or address* or halt* or stop* or abat* or remission* or moderat* or modif* or control* or zero or diminish* or decreas* or eliminat* or elevat* or emission* or rise* or increas* or declin* or sink* or sequester* or neutral or offset or storage or store or storing or captur* or perform* or efficien* or conservation or remov*)))

78,512

S10 (AB(emission* N6 (elevat* or rise or increas* or decreas* or reduc* or offset*))) OR (TI(emission* N6 (elevat* or rise or increas* or decreas* or reduc* or offset*))) 36,555

S11 (TI “tipping point#”) or (AB “tipping point#”) 510

S12 (TI (carbon N4 footprint#)) or (AB (carbon N4 footprint#)) 2,770

S13 (AB(de-carbon or de-carboni* or decarbon or decarboni*)) OR (TI(de-carbon or de-carboni* or decarbon or decarboni*)) 2,654

S14 (AB((energy* or fuel*) N4 (green or efficien* or reduc* or renewable* or save* or saving or clean* or transition*))) OR (TI((energy* or fuel*) N4 (green or efficien* or reduc* or renewable* or save* or saving or clean* or transition*))) 62,050

S15 (AB((“fossil fuel*” or coal or oil or gas or carbon) N4 (“phas* out” or exit* or transition* or transform* or clos* or shift* or decommission* or “shut* down” or displac* or low* or neutral* or sustainab*))) OR (TI((“fossil fuel*” or coal or oil or gas or carbon) N4 (“phas* out” or exit* or transition* or transform* or clos* or shift* or decommission* or “shut* down” or displac* or low* or neutral* or sustainab*))) 19,187

S16 (TI (net-zero or netzero)) OR (AB (net-zero or netzero)) 1,119 S17 (TI “just transition*”) OR (AB “just transition*”) 183

S18 (TI BECCS) OR (AB BECCS) 160

S19 (TI petagram*) or (AB petagram*) 14

S20 (MH “Wildfires”) 2,410

S21 (MH “Natural Disasters”) 1,915

S22 (TI ((rain* or precipitation) N5 (extreme or elevate# or elevating or irregular or intense or reduced or pattern* or anomal* or deficit* or surplus* or shortfall*))) or (AB ((rain* or precipitation) N5 (extreme or elevate# or elevating or irregular or intense or reduced or pattern* or anomal* or deficit* or surplus* or shortfall*))) 6,754

S23 (TI (((fire# or burnt or inferno or conflagration) N3 (change* or variab* or extreme or expand or expansion or intense or elevate# or increas*or catastroph* or emergenc* or shift* or dynamic* or frequen* or more or forest* or landscape# or wildland*)) or wildfire* or bushfire* or megafire*) or (AB (((fire# or burnt or inferno or conflagration) N3 (change* or variab* or extreme or expand or expansion or intense or elevate# or increas*or catastroph* or emergenc* or shift* or dynamic* or frequen* or more or forest* or landscape# or wildland*)) or wildfire* or bushfire* or megafire*) 9,738

S24 (TI (flood* or (water N3 (stress or deficit or scarce or scarcit*)) or drought# or (climat* N3 (wet* or dry*)))) or (AB (flood* or (water N3 (stress or deficit or scarce or scarcit*)) or drought# or (climat* N3 (wet* or dry*)))) 30,591

S25 (TI (habitat N3 (loss or fragment* or disturb* or alter or altered or destruct*))) or (AB (habitat N3 (loss or fragment* or disturb* or alter or altered or destruct*))) 7,904

S26 (TI (deforestation or reforestation or “slash-and-burn” or desertificat* or (forest* adj2 (degradat* or conservat* or declin* or decreas*)))) or (AB (deforestation or reforestation or “slash-and-burn” or desertificat* or (forest* adj2 (degradat* or conservat* or declin* or decreas*)))) 7,901

S27 S1 OR S2 OR S3 OR S4 OR S5 OR S6 OR S7 OR S8 OR S9 OR S10 OR S11 OR S12 OR S13 OR S14 OR S15 OR S16 OR S17 OR S18 OR S19 OR S20 OR S21 OR S22 OR S23 OR S24 OR S25 OR S26 259,552

S28 (MH “Diarrhea”) 166

S29 (TI (diarrhoea* or diarrhea*)) or (AB (diarrhoea* or diarrhea*)) 463

S30 (TI ((water-borne or waterborne) N2 (disease* or infect* or illness* or pathogen*))) or (AB ((water-borne or waterborne) N2 (disease* or infect* or illness* or pathogen*))) 516

S31 (MH “Gastroenteritis”) 72

S32 (MH “Intestinal Diseases, Parasitic”) 0

S33 (TI (gastroenteritis or ((infect* or illness* or pathogen* or microbio* or bacteri* or virus* or viral or protozoa*) N2 (intestin* or enteric* or gastro* or bowel)))) or (AB (gastroenteritis or ((infect* or illness* or pathogen* or microbio* or bacteri* or virus* or viral or protozoa*) N2 (intestin* or enteric* or gastro* or bowel)))) 1,179

S34 (MH “Campylobacter”) 155

S35 (MH “Campylobacter Infections”) 38

S36 (TI campylobacter*) or (AB campylobacter*) 259

S37 (MH “Giardiasis”) 15

S38 (MH “Giardia Lamblia”) 36

S39 (TI (giardiasis or giardiases or giardia)) or (AB (giardiasis or giardiases or giardia)) 246

S40 (MH “Cryptosporidiosis”) 53

S41 (MH “Cryptosporidium”) 301

S42 (TI (cryptosporidiosis or cryptosporidioses or cryptosporidium)) or (AB (cryptosporidiosis or cryptosporidioses or cryptosporidium)) 445

S43 (MH “Rotaviruses”) OR (MH “Rotavirus Infections”) 35

S44 (TI rotavirus) or (AB rotavirus) 100

S45 S28 OR S29 OR S30 OR S31 OR S32 OR S33 OR S34 OR S35 OR S36 OR S37 OR S38 OR S39 OR S40 OR S41 OR S42 OR S43 OR S44 2,680

S46 (MH “Risk Assessment”) 25,163 S47 (MH “Risk Management”) 2,000 S48 (MH “Attributable Risk”) 42

S49 (MH “Adaptation, Physiological+”) 0

S50 (MH “Adaptation, Psychological”) 0

S51 (MH “Vulnerability”) 2,745

S52 (AB (adaptation* or risk or vulnerable or vulnerabilit* or sustainab* or resilient or resilienc*)) or (TI (adaptation* or risk or vulnerable or vulnerabilit* or sustainab* or resilient or resilienc*)) 202,794

S53 S46 OR S47 OR S48 OR S49 OR S50 OR S51 OR S52 207,540 S54 S27 AND S45 AND S53 156

Clarivate Analytics Web of Science Core Content

Database name Science Citation Index Expanded (SCI-EXPANDED) Social Sciences Citation Index (SSCI)

Arts & Humanities Citation Index (SSCI)

Conference Proceedings Citation Index-Science (CPCI-S)

Conference Proceedings Citation Index-Social Science & Humanities (CPCI-SSH) Emerging Sources Citation Index (ESCI)

Database platform Clarivate Analytics Web of Science Dates of database coverage

SCI-EXPANDED, 1970-present

SSCI, 1970-present AHCI, 1975-present CPCI-S, 1990-present CPCI-SSH, 1990-present ESCI, 2019-present

Data updated 2024-11-12

Date searched 2024-11-15

Number of results 1065

EndNote deduplication order 7

Results after duplications removed 328

Notes

Two-letter codes at the beginning of search lines designate the fields to search. Fields codes used are: TI: title

AB: abstract

NEAR/n searches for words within n words of each other.

* is used for truncation of words.

$ finds 0 or 1 character in that placeholder (optional wildcard). All databases are searched simultaneously.

1: TI=(“climat*” NEAR/4 (“change$” or “changing” or “variab*” or “disrupt*” or “hazard*” or “impact*” or “vulnerab*” or “function*” or “extreme” or “catastroph*” or “emergenc*” or “shift*” or “dynamic*” or “sensitiv*”)) OR AB=(“climat*” NEAR/4 (“change$” or “changing” or “variab*” or “disrupt*” or “hazard*” or “impact*” or “vulnerab*” or “function*” or “extreme” or “catastroph*” or “emergenc*” or “shift*” or “dynamic*” or “sensitiv*”)) Results: 421210

2: TI=((“global” or “planet*” or “world” or “earth” or “worldwide” or “world-wide”) NEAR/3 (“co2” or “carbon” or “methane” or “ozone” or “nitrous oxide”)) OR AB=((“global” or “planet*” or “world” or “earth” or “worldwide” or “world-wide”) NEAR/3 (“co2” or “carbon” or “methane” or “ozone” or “nitrous oxide”)) Results: 37749

3: TI=((“greenhouse” or “green-house”) NEAR/3 (“gas*” or “effect*”)) OR AB=((“greenhouse” or “green-house”) NEAR/3 (“gas*” or “effect*”)) Results: 102281

4: TI=((“co2” or “carbon” or “GHG” or “CH4” or “methane” or “N2O” or “nitro* oxide*” or “non-methane volatile organic compound*” or “NMVOC*” or “sulphur dioxide*” or “ozone” or “O3” or “fluorinated gase*” or “F-Gas*” or “hydrofluorocarbon*” or “HFCs” or “perfluorocarbon*” or “PFCs” or “sulfur hexafluoride*” or “sulphur hexafluoride*” or “SF6” or “chlorofluorocarbon*” or “CFCs” or “hydrochlorofluorocarbon*” or “HCFCs” or “halos” or “air pollut*” or “aerosol*” or ((“short-lived” or “shortlived”) NEAR/4 “pollutant*”) or “SLCP*”) NEAR/4 (“mitigat*” or “reduc*” or “save*” or “saving” or “tackl*” or “address*” or “halt*” or “stop*” or “abat*” or “remission*” or “moderat*” or “modif*” or “control*” or “zero” or “diminish*” or “decreas*” or “eliminat*” or “elevat*” or “emission*” or “rise*” or “increas*” or “declin*” or “sink*” or “sequester*” or “neutral” or “offset” or “storage” or “store” or “storing” or “captur*” or “perform*” or “efficien*” or “conservation” or “remov*”)) OR AB=((“co2” or “carbon” or “GHG” or “CH4” or “methane” or “N2O” or “nitro* oxide*” or “non-methane volatile organic compound*” or “NMVOC*” or “sulphur dioxide*” or “ozone” or “O3” or “fluorinated gase*” or “F-Gas*” or “hydrofluorocarbon*” or “HFCs” or “perfluorocarbon*” or “PFCs” or “sulfur hexafluoride*” or “sulphur hexafluoride*” or “SF6” or “chlorofluorocarbon*” or “CFCs” or “hydrochlorofluorocarbon*” or “HCFCs” or “halos” or “air pollut*” or “aerosol*” or ((“short-lived” or “shortlived”) NEAR/4 “pollutant*”) or “SLCP*”) NEAR/4 (“mitigat*” or “reduc*” or “save*” or “saving” or “tackl*” or “address*” or “halt*” or “stop*” or “abat*” or “remission*” or “moderat*” or “modif*” or “control*” or “zero” or “diminish*” or “decreas*” or “eliminat*” or “elevat*” or “emission*” or “rise*” or “increas*” or “declin*” or “sink*” or “sequester*” or “neutral” or “offset” or “storage” or “store” or “storing” or “captur*” or “perform*” or “efficien*” or “conservation” or “remov*”)) Results: 705783

5: TI=(“emission*” NEAR/6 (“elevat*” or “rise” or “increas*” or “decreas*” or “reduc*” or “offset*”)) OR AB=(“emission*” NEAR/6 (“elevat*” or “rise” or “increas*” or “decreas*” or “reduc*” or “offset*”)) Results: 221294

6: TI=“tipping point$” or AB=“tipping point$” Results: 4555

7: TI=(“carbon” NEAR/4 “footprint$”) OR AB=(“carbon” NEAR/4 “footprint$”) Results: 18053

8: TI=(“de-carbon” or “de-carboni*” or “decarbon” or “decarboni*”) or AB=(“de-carbon” or “de-carboni*” or “decarbon” or “decarboni*”) Results: 16845

9: TI=((“energy*” or “fuel*”) NEAR/4 (“green” or “efficien*” or “reduc*” or “renewable*” or “save*” or “saving” or “clean*” or “transition*”)) or AB=((“energy*” or “fuel*”) NEAR/4 (“green” or “efficien*” or “reduc*” or “renewable*” or “save*” or “saving” or “clean*” or “transition*”))

Results: 714405

10: TI=((“fossil fuel*” or “coal” or “oil” or “gas” or “carbon”) NEAR/4 (“phas* out” or “exit*” or “transition*” or “transform*” or “clos*” or “shift*” or “decommission*” or “shut* down” or “displac*” or “low*” or “neutral*” or “sustainab*”)) or AB=((“fossil fuel*” or “coal” or “oil” or “gas” or “carbon”) NEAR/4 (“phas* out” or “exit*” or “transition*” or “transform*” or “clos*” or “shift*” or “decommission*” or “shut* down” or “displac*” or “low*” or “neutral*” or “sustainab*”)) Results: 282794

11: TI=(“net-zero” or “netzero”) or AB=(“net-zero” or “netzero”) Results: 7854 12: TI=“just transition*” or AB=“just transition*” Results: 914

13: TI=BECCS or AB=BECCS Results: 515

14: TI=pentagram* or AB=pentagram* Results: 278

15: TI=((“rain*” or “precipitation”) NEAR/5 (“extreme” or “elevate#” or “elevating” or “irregular” or “intense” or “reduced” or “pattern*” or “anomal*” or “deficit*” or “surplus*” or “shortfall*”)) or AB=((“rain*” or “precipitation”) NEAR/5 (“extreme” or “elevate#” or “elevating” or “irregular” or “intense” or “reduced” or “pattern*” or “anomal*” or “deficit*” or “surplus*” or “shortfall*”))

Results: 54524

16: TI=(((“fire#” or “burnt” or “inferno” or “conflagration”) NEAR/3 (“change*” or “variab*” or “extreme” or “expand” or “expansion” or “intense” or “elevate#” or “increas*”or “catastroph*” or “emergenc*” or “shift*” or “dynamic*” or “frequen*” or “more” or “forest*” or “landscape#” or “wildland*”)) or “wildfire*” or “bushfire*” or “megafire*”) or AB=(((“fire#” or “burnt” or “inferno” or “conflagration”) NEAR/3 (“change*” or “variab*” or “extreme” or “expand” or “expansion” or “intense” or “elevate#” or “increas*”or “catastroph*” or “emergenc*” or “shift*” or “dynamic*” or “frequen*” or “more” or “forest*” or “landscape#” or “wildland*”)) or “wildfire*” or “bushfire*” or “megafire*”)

Results: 52243

17: TI=(“flood*” or (“water” NEAR/3 (“stress” or “deficit” or “scarce” or “scarcit*”)) or “drought#” or (“climat*” NEAR/3 (“wet*” or “dry*”))) or AB=(“flood*” or (“water” NEAR/3 (“stress” or “deficit” or “scarce” or “scarcit*”)) or “drought#” or (“climat*” NEAR/3 (“wet*” or “dry*”)))

Results: 362085

18: TI=(“habitat” NEAR/3 (“loss” or “fragment*” or “disturb*” or “alter” or “altered” or “destruct*”)) or AB=(“habitat” NEAR/3 (“loss” or “fragment*” or “disturb*” or “alter” or “altered” or “destruct*”)) Results: 26652

19: TI=(“deforestation” or “reforestation” or “slash-and-burn” or “desertificat*” or (“forest*” NEAR/3 (“degradat*” or “conservat*” or “declin*” or “decreas*”))) or AB=(“deforestation” or “reforestation” or “slash-and-burn” or “desertificat*” or (“forest*” NEAR/3 (“degradat*” or “conservat*” or “declin*” or “decreas*”))) Results: 54392

20: #19 OR #18 OR #17 OR #16 OR #15 OR #14 OR #13 OR #12 OR #11 OR #10 OR #9 OR #8 OR #7 OR #6 OR #5 OR #4 OR #3 OR #2 OR

#1 Results: 2411546

21: TI=(“diarrhoea*” or “diarrhea*”) or AB=(“diarrhoea*” or “diarrhea*”) Results: 119690

22: TI=((“faecal” or “fecal” or “faeces” or “feces” or “stool#”) NEAR/3 (“consisten*” or “liquid*” or “water*”)) or AB=((“faecal” or “fecal” or “faeces” or “feces” or “stool#”) NEAR/3 (“consisten*” or “liquid*” or “water*”)) Results: 9317

23: TI=((“water-borne” or “waterborne”) NEAR/2 (“disease*” or “infect*” or “illness*” or “pathogen*”)) or AB=((“water-borne” or “waterborne”) NEAR/2 (“disease*” or “infect*” or “illness*” or “pathogen*”)) Results: 4716

24: TI=(“gastroenteritis” or ((“infect*” or “illness*” or “pathogen*” or “microbio*” or “bacteri*” or “virus*” or “viral” or “protozoa*”) NEAR/2 (“intestin*” or “enteric*” or “gastro*” or “bowel”))) or AB=(“gastroenteritis” or ((“infect*” or “illness*” or “pathogen*” or “microbio*” or “bacteri*” or “virus*” or “viral” or “protozoa*”) NEAR/2 (“intestin*” or “enteric*” or “gastro*” or “bowel”)))

Results: 104463

25: TI=“campylobacter*” or AB=“campylobacter*” Results: 22392

26: TI=(“giardiasis” or “giardiases” or “giardia”) or AB=(“giardiasis” or “giardiases” or “giardia”) Results: 10780

27: TI=(“cryptosporidiosis” or “cryptosporidioses” or “cryptosporidium”) or AB=(“cryptosporidiosis” or “cryptosporidioses” or “cryptosporidium”) Results: 12285

28: TI=“rotavirus” or AB=“rotavirus” Results: 16747

29: #28 OR #27 OR #26 OR #25 OR #24 OR #23 OR #22 OR #21 Results: 259590

30: TI=(“adaptation*” or “risk” or “vulnerable” or “vulnerabilit*” or “sustainab*” or “resilient” or “resilienc*”) or AB=(“adaptation*” or “risk” or “vulnerable” or “vulnerabilit*” or “sustainab*” or “resilient” or “resilienc*”) Results: 5130547

31: #30 AND #29 AND #20 Results: 1065

Clarivate Analytics Web of Science BIOSIS Citation Index Database name BIOSIS Citation Index

Database platform Clarivate Analytics Web of Science

Dates of database coverage Complete database. Data updated 2024-11-12 Date searched 2024-11-15

Number of results 627

EndNote deduplication order 8

Results after duplications removed 38

Notes

Two-letter codes at the beginning of search lines designate the fields to search. Fields codes used are: TI: title

AB: abstract

NEAR/n searches for words within n words of each other.

* is used for truncation of words.

$ finds 0 or 1 character in that placeholder (optional wildcard).

1: TI=(“climat*” NEAR/4 (“change$” or “changing” or “variab*” or “disrupt*” or “hazard*” or “impact*” or “vulnerab*” or “function*” or “extreme” or “catastroph*” or “emergenc*” or “shift*” or “dynamic*” or “sensitiv*”)) OR AB=(“climat*” NEAR/4 (“change$” or “changing” or “variab*” or “disrupt*” or “hazard*” or “impact*” or “vulnerab*” or “function*” or “extreme” or “catastroph*” or “emergenc*” or “shift*” or “dynamic*” or “sensitiv*”)) Results: 179850

2: TI=((“global” or “planet*” or “world” or “earth” or “worldwide” or “world-wide”) NEAR/3 (“co2” or “carbon” or “methane” or “ozone” or “nitrous oxide”)) OR AB=((“global” or “planet*” or “world” or “earth” or “worldwide” or “world-wide”) NEAR/3 (“co2” or “carbon” or “methane” or “ozone” or “nitrous oxide”)) Results: 13784

3: TI=((“greenhouse” or “green-house”) NEAR/3 (“gas*” or “effect*”)) OR AB=((“greenhouse” or “green-house”) NEAR/3 (“gas*” or “effect*”)) Results: 29606

4: TI=((“co2” or “carbon” or “GHG” or “CH4” or “methane” or “N2O” or “nitro* oxide*” or “non-methane volatile organic compound*” or “NMVOC*” or “sulphur dioxide*” or “ozone” or “O3” or “fluorinated gase*” or “F-Gas*” or “hydrofluorocarbon*” or “HFCs” or “perfluorocarbon*” or “PFCs” or “sulfur hexafluoride*” or “sulphur hexafluoride*” or “SF6” or “chlorofluorocarbon*” or “CFCs” or “hydrochlorofluorocarbon*” or “HCFCs” or “halos” or “air pollut*” or “aerosol*” or ((“short-lived” or “shortlived”) NEAR/4 “pollutant*”) or “SLCP*”) NEAR/4 (“mitigat*” or “reduc*” or “save*” or “saving” or “tackl*” or “address*” or “halt*” or “stop*” or “abat*” or “remission*” or “moderat*” or “modif*” or “control*” or “zero” or “diminish*” or “decreas*” or “eliminat*” or “elevat*” or “emission*” or “rise*” or “increas*” or “declin*” or “sink*” or “sequester*” or “neutral” or “offset” or “storage” or “store” or “storing” or “captur*” or “perform*” or “efficien*” or “conservation” or “remov*”)) OR AB=((“co2” or “carbon” or “GHG” or “CH4” or “methane” or “N2O” or “nitro* oxide*” or “non-methane volatile organic compound*” or “NMVOC*” or “sulphur dioxide*” or “ozone” or “O3” or “fluorinated gase*” or “F-Gas*” or “hydrofluorocarbon*” or “HFCs” or “perfluorocarbon*” or “PFCs” or “sulfur hexafluoride*” or “sulphur hexafluoride*” or “SF6” or “chlorofluorocarbon*” or “CFCs” or “hydrochlorofluorocarbon*” or “HCFCs” or “halos” or “air pollut*” or “aerosol*” or ((“short-lived” or “shortlived”) NEAR/4 “pollutant*”) or “SLCP*”) NEAR/4 (“mitigat*” or “reduc*” or “save*” or “saving” or “tackl*” or “address*” or “halt*” or “stop*” or “abat*” or “remission*” or “moderat*” or “modif*” or “control*” or “zero” or “diminish*” or “decreas*” or “eliminat*” or “elevat*” or “emission*” or “rise*” or “increas*” or “declin*” or “sink*” or “sequester*” or “neutral” or “offset” or “storage” or “store” or “storing” or “captur*” or “perform*” or “efficien*” or “conservation” or “remov*”)) Results: 217707

5: TI=(“emission*” NEAR/6 (“elevat*” or “rise” or “increas*” or “decreas*” or “reduc*” or “offset*”)) OR AB=(“emission*” NEAR/6 (“elevat*” or “rise” or “increas*” or “decreas*” or “reduc*” or “offset*”)) Results: 48769

6: TI=“tipping point$” or AB=“tipping point$” Results: 1585

7: TI=(“carbon” NEAR/4 “footprint$”) OR AB=(“carbon” NEAR/4 “footprint$”) Results: 3275

8: TI=(“de-carbon” or “de-carboni*” or “decarbon” or “decarboni*”) or AB=(“de-carbon” or “de-carboni*” or “decarbon” or “decarboni*”) Results: 1177

9: TI=((“energy*” or “fuel*”) NEAR/4 (“green” or “efficien*” or “reduc*” or “renewable*” or “save*” or “saving” or “clean*” or “transition*”)) or AB=((“energy*” or “fuel*”) NEAR/4 (“green” or “efficien*” or “reduc*” or “renewable*” or “save*” or “saving” or “clean*” or “transition*”))

Results: 72598

10: TI=((“fossil fuel*” or “coal” or “oil” or “gas” or “carbon”) NEAR/4 (“phas* out” or “exit*” or “transition*” or “transform*” or “clos*” or “shift*” or “decommission*” or “shut* down” or “displac*” or “low*” or “neutral*” or “sustainab*”)) or AB=((“fossil fuel*” or “coal” or “oil” or “gas” or “carbon”) NEAR/4 (“phas* out” or “exit*” or “transition*” or “transform*” or “clos*” or “shift*” or “decommission*” or “shut* down” or “displac*” or “low*” or “neutral*” or “sustainab*”)) Results: 52836

11: TI=(“net-zero” or “netzero”) or AB=(“net-zero” or “netzero”) Results: 727 12: TI=“just transition*” or AB=“just transition*” Results: 44

13: TI=BECCS or AB=BECCS Results: 99

14: TI=pentagram* or AB=pentagram* Results: 42

15: TI=((“rain*” or “precipitation”) NEAR/5 (“extreme” or “elevate#” or “elevating” or “irregular” or “intense” or “reduced” or “pattern*” or “anomal*” or “deficit*” or “surplus*” or “shortfall*”)) or AB=((“rain*” or “precipitation”) NEAR/5 (“extreme” or “elevate#” or “elevating” or “irregular” or “intense” or “reduced” or “pattern*” or “anomal*” or “deficit*” or “surplus*” or “shortfall*”))

Results: 18816

16: TI=(((“fire#” or “burnt” or “inferno” or “conflagration”) NEAR/3 (“change*” or “variab*” or “extreme” or “expand” or “expansion” or “intense” or “elevate#” or “increas*”or “catastroph*” or “emergenc*” or “shift*” or “dynamic*” or “frequen*” or “more” or “forest*” or “landscape#” or “wildland*”)) or “wildfire*” or “bushfire*” or “megafire*”) or AB=(((“fire#” or “burnt” or “inferno” or “conflagration”) NEAR/3 (“change*” or “variab*” or “extreme” or “expand” or “expansion” or “intense” or “elevate#” or “increas*”or “catastroph*” or “emergenc*” or “shift*” or “dynamic*” or “frequen*” or “more” or “forest*” or “landscape#” or “wildland*”)) or “wildfire*” or “bushfire*” or “megafire*”)

Results: 25381

17: TI=(“flood*” or (“water” NEAR/3 (“stress” or “deficit” or “scarce” or “scarcit*”)) or “drought#” or (“climat*” NEAR/3 (“wet*” or “dry*”))) or AB=(“flood*” or (“water” NEAR/3 (“stress” or “deficit” or “scarce” or “scarcit*”)) or “drought#” or (“climat*” NEAR/3 (“wet*” or “dry*”)))

Results: 184041

18: TI=(“habitat” NEAR/3 (“loss” or “fragment*” or “disturb*” or “alter” or “altered” or “destruct*”)) or AB=(“habitat” NEAR/3 (“loss” or “fragment*” or “disturb*” or “alter” or “altered” or “destruct*”)) Results: 26103

19: TI=(“deforestation” or “reforestation” or “slash-and-burn” or “desertificat*” or (“forest*” NEAR/3 (“degradat*” or “conservat*” or “declin*” or “decreas*”))) or AB=(“deforestation” or “reforestation” or “slash-and-burn” or “desertificat*” or (“forest*” NEAR/3 (“degradat*” or “conservat*” or “declin*” or “decreas*”))) Results: 35712

20: #19 OR #18 OR #17 OR #16 OR #15 OR #14 OR #13 OR #12 OR #11 OR #10 OR #9 OR #8 OR #7 OR #6 OR #5 OR #4 OR #3 OR #2 OR

#1 Results: 722520

21: TI=(“diarrhoea*” or “diarrhea*”) or AB=(“diarrhoea*” or “diarrhea*”) Results: 89366

22: TI=((“faecal” or “fecal” or “faeces” or “feces” or “stool#”) NEAR/3 (“consisten*” or “liquid*” or “water*”)) or AB=((“faecal” or “fecal” or “faeces” or “feces” or “stool#”) NEAR/3 (“consisten*” or “liquid*” or “water*”)) Results: 8077

23: TI=((“water-borne” or “waterborne”) NEAR/2 (“disease*” or “infect*” or “illness*” or “pathogen*”)) or AB=((“water-borne” or “waterborne”) NEAR/2 (“disease*” or “infect*” or “illness*” or “pathogen*”)) Results: 3260

24: TI=(“gastroenteritis” or ((“infect*” or “illness*” or “pathogen*” or “microbio*” or “bacteri*” or “virus*” or “viral” or “protozoa*”) NEAR/2 (“intestin*” or “enteric*” or “gastro*” or “bowel”))) or AB=(“gastroenteritis” or ((“infect*” or “illness*” or “pathogen*” or “microbio*” or “bacteri*” or “virus*” or “viral” or “protozoa*”) NEAR/2 (“intestin*” or “enteric*” or “gastro*” or “bowel”)))

Results: 83206

25: TI=“campylobacter*” or AB=“campylobacter*” Results: 19879

26: TI=(“giardiasis” or “giardiases” or “giardia”) or AB=(“giardiasis” or “giardiases” or “giardia”) Results: 9846

27: TI=(“cryptosporidiosis” or “cryptosporidioses” or “cryptosporidium”) or AB=(“cryptosporidiosis” or “cryptosporidioses” or “cryptosporidium”) Results: 10864

28: TI=“rotavirus” or AB=“rotavirus” Results: 13977

29: #28 OR #27 OR #26 OR #25 OR #24 OR #23 OR #22 OR #21 Results: 204713

30: TI=(“adaptation*” or “risk” or “vulnerable” or “vulnerabilit*” or “sustainab*” or “resilient” or “resilienc*”) or AB=(“adaptation*” or “risk” or “vulnerable” or “vulnerabilit*” or “sustainab*” or “resilient” or “resilienc*”) Results: 2220911

31: #30 AND #29 AND #20 Results: 627

Elsevier Scopus

Database name Scopus

Database platform Elsevier scopus.com

Dates of database coverage: Complete database to search date Date searched 2024-11-15

Number of results 1537

EndNote deduplication order 9

Results after duplications removed 388

Notes

All searches run on the title and abstract.

W/n searches for words within n words of each other.

* is used for truncation of words.

TITLE-ABS((“climat*” W/4 (“change*” or “changing” or “variab*” or “disrupt*” or “hazard*” or “impact*” or “vulnerab*” or “function*” or “extreme” or “catastroph*” or “emergenc*” or “shift*” or “dynamic*” or “sensitiv*”)) or ((“global” or “planet*” or “world” or “earth” or “worldwide” or “world-wide”) W/3 (“co2” or “carbon” or “methane” or “ozone” or “nitrous oxide”)) or ((“greenhouse” or “green-house”) W/3 (“gas*” or “effect*”)) OR ((“co2” or “carbon” or “GHG” or “CH4” or “methane” or “N2O” or “nitro* oxide*” or “non-methane volatile organic compound*” or “NMVOC*” or “sulphur dioxide*” or “ozone” or “O3” or “fluorinated gase*” or “F-Gas*” or “hydrofluorocarbon*” or “HFCs” or “perfluorocarbon*” or “PFCs” or “sulfur hexafluoride*” or “sulphur hexafluoride*” or “SF6” or “chlorofluorocarbon*” or “CFCs” or “hydrochlorofluorocarbon*” or “HCFCs” or “halos” or “air pollut*” or “aerosol*” or ((“short-lived” or “shortlived”) W/4 “pollutant*”) or “SLCP*”) W/4 (“mitigat*” or “reduc*” or “save*” or “saving” or “tackl*” or “address*” or “halt*” or “stop*” or “abat*” or “remission*” or “moderat*” or “modif*” or “control*” or “zero” or “diminish*” or “decreas*” or “eliminat*” or “elevat*” or “emission*” or “rise*” or “increas*” or “declin*” or “sink*” or “sequester*” or “neutral” or “offset” or “storage” or “store” or “storing” or “captur*” or “perform*” or “efficien*” or “conservation” or “remov*”)) or (“emission*” W/6 (“elevat*” or “rise” or “increas*” or “decreas*” or “reduc*” or “offset*”)) or “tipping point$” or (“carbon” W/4 “footprint$”) or (“de-carbon” or “de-carboni*” or “decarbon” or “decarboni*”) or ((“energy*” or “fuel*”) W/4 (“green” or “efficien*” or “reduc*” or “renewable*” or “save*” or “saving” or “clean*” or “transition*”)) or ((“fossil fuel*” or “coal” or “oil” or “gas” or “carbon”) W/4 (“phas* out” or “exit*” or “transition*” or “transform*” or “clos*” or “shift*” or “decommission*” or “shut* down” or “displac*” or “low*” or “neutral*” or “sustainab*”)) or (“net-zero” or “netzero”) or “just transition*” or ((“rain*” or “precipitation”) W/5 (“extreme” or “elevate*” or “elevating” or “irregular” or “intense” or “reduced” or “pattern*” or “anomal*” or “deficit*” or “surplus*” or “shortfall*”)) or ((“fire*” or “burnt” or “inferno” or “conflagration”) W/3 (“change*” or “variab*” or “extreme” or “expand” or “expansion” or “intense” or “elevate*” or “increas*”or “catastroph*” or “emergenc*” or “shift*” or “dynamic*” or “frequen*” or “more” or “forest*” or “landscape*” or “wildland*”)) or “wildfire*” or “bushfire*” or “megafire*” or “flood*” or (“water” W/3 (“stress” or “deficit” or “scarce” or “scarcit*”)) or “drought*” or (“climat*” W/3 (“wet*” or “dry*”)) or (“habitat” W/3 (“loss” or “fragment*” or “disturb*” or “alter” or “altered” or “destruct*”)) or “deforestation” or “reforestation” or “slash-and-burn” or “desertificat*” or (“forest*” W/3 (“degradat*” or “conservat*” or “declin*” or “decreas*”))) AND TITLE-ABS(“diarrhoea*” or “diarrhea*” or ((“faecal” or “fecal” or “faeces” or “feces” or “stool*”) W/3 (“consisten*” or “liquid*” or “water*”)) or ((“water-borne” or “waterborne”) W/2 (“disease*” or “infect*” or “illness*” or “pathogen*”)) or “gastroenteritis” or ((“infect*” or “illness*” or “pathogen*” or “microbio*” or “bacteri*” or “virus*” or “viral” or “protozoa*”) W/2 (“intestin*” or “enteric*” or “gastro*” or “bowel”)) or “campylobacter*” or “giardiasis” or “giardiases” or “giardia” or “cryptosporidiosis” or “cryptosporidioses” or “cryptosporidium” or “rotavirus”) AND TITLE-ABS(“adaptation*” or “risk” or “vulnerable” or “vulnerabilit*” or “sustainab*” or “resilient” or “resilienc*”)

## Appendix 3: Data extraction table

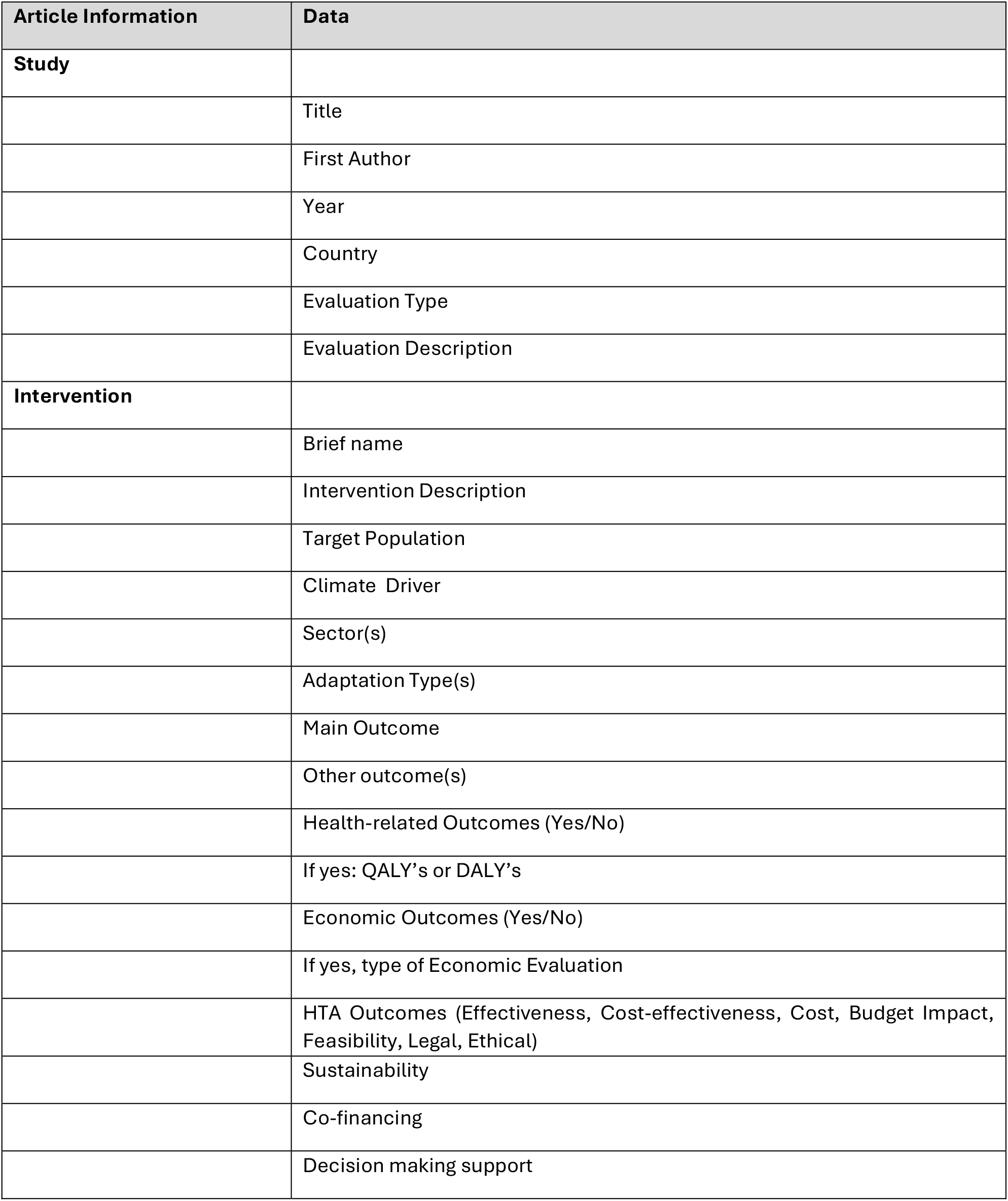

| Article Information | Data |
| --- | --- |
| **Study** |  |
|  | Title |
|  | First Author |
|  | Year |
|  | Country |
|  | Evaluation Type |
|  | Evaluation Description |
| **Intervention** |  |
|  | Brief name |
|  | Intervention Description |
|  | Target Population |
|  | Climate Driver |
|  | Sector(s) |
|  | Adaptation Type(s) |
|  | Main Outcome |
|  | Other outcome(s) |
|  | Health-related Outcomes (Yes/No) |
|  | If yes: QALY's or DALY's |
|  | Economic Outcomes (Yes/No) |
|  | If yes, type of Economic Evaluation |
|  | HTA Outcomes (Effectiveness, Cost-effectiveness, Cost, Budget Impact, Feasibility, Legal, Ethical) |
|  | Sustainability |
|  | Co-financing |
|  | Decision making support |

